# Temporal trends in test-seeking behaviour during the COVID-19 pandemic

**DOI:** 10.1101/2024.06.06.24308566

**Authors:** Oliver Eales, Mingmei Teo, David J. Price, Tianxiao Hao, Gerard E. Ryan, Katharine L. Senior, Sandra Carlson, Craig Dalton, Peter Dawson, Nick Golding, James M. McCaw, Freya M. Shearer

**Author notes:** Corresponding author: Freya Shearer.

## Abstract

**Background:** During the COVID-19 pandemic, many countries implemented mass community testing programs, where individuals would seek tests due to (primarily) the onset of symptoms. The cases recorded by mass testing programs represent only a fraction of infected individuals, and depend on how many people seek testing. If test-seeking behaviour exhibits heterogeneities or changes over time, and this is not accounted for when analysing case data, then inferred epidemic dynamics used to inform public health decision-making can be biased.

**Methods:** Here we describe temporal trends in COVID-19 test-seeking behaviour in Australia by symptoms, age group, test type, and jurisdiction from November 2021–September 2023. We use data from two surveillance systems: a weekly nationwide behavioural survey (NBS), established by the Australian Government to monitor a range of behavioural responses to COVID-19; and Australia’s FluTracking system, a ‘participatory surveillance system’ designed for monitoring influenza-like illness and health-care seeking behaviour, which was adapted in early 2020 to include questions relevant to COVID-19.

**Results:** We found that peaks in test-seeking behaviour generally aligned with peaks in the rate of reported cases. Test-seeking behaviour rapidly increased in early-2022 coinciding with greater availability of rapid antigen tests. There were heterogeneities in test-seeking behaviour by jurisdiction and age-group, which were dynamic through time. Test-seeking behaviour was lowest in older individuals (60+ years) until July 2022, after which there was greater homogeneity across age-groups. Test-seeking behaviour was highest in the Australian Capital Territory and Tasmania and consistently lowest in Queensland. Over the course of the study test-seeking behaviour was highest in individuals who reported symptoms more predictive of COVID-19 infection. There was a greater probability of seeking a test for individuals in FluTracking compared to the NBS, suggesting that participatory surveillance systems such as FluTracking may include a health-conscious subset of the population.

**Conclusions:** Our findings demonstrate the dynamism of test-seeking behaviour, highlighting the importance of the continued collection of behavioural data through dedicated surveillance systems.

## Introduction

How do infectious disease surveillance systems identify infected individuals? Understanding the processes leading to a case of disease being recorded by a surveillance system is required to appropriately interpret and analyse surveillance data. These processes are influenced by a myriad of factors including public health interventions, the behaviour of both infected individuals and health practitioners, data collection practices, and provision of laboratory services.

During the COVID-19 pandemic, many countries implemented mass community testing programs. The case-based surveillance data generated by mass testing were often used by epidemiologists to understand, monitor, and predict epidemic activity [1] including the potential exceedance of hospital capacity [2]. However, such analyses are fraught with challenges since the relationship between the time-series of cases and the underlying infection dynamics is difficult to characterise [3]. The COVID-19 cases recorded by mass testing programs represent only a fraction of individuals infected with SARS-CoV-2 [4] which depends on how many people seek and access testing. The daily number of COVID-19 tests was routinely reported in many countries and shown to change (dramatically) over time. However, the extent to which testing rates are driven by changes in the infection incidence cannot be determined without data on test-seeking behaviour.

Standard influenza surveillance systems — which rely on infected individuals seeking care at health facilities — are sometimes supplemented by data on healthcare-seeking behaviour collected through targeted population surveys and “participatory surveillance systems”. The latter conduct regular surveys of self-reporting community volunteer cohorts. Participatory surveillance systems operate around the world, including ‘FluTracking’ in Australia (since 2006), New Zealand (since 2018), Hong Kong (since 2020) and Argentina (since 2022) [5], ‘Influenzanet’ in many European countries (since 2003) [6] and ‘Outbreaks Near Me’ (previously called ‘Flu Near You’) in North America (since 2011) [7].

Analyses of these data have shown that healthcare-seeking behaviour is dynamic through time [8,9], can change rapidly in response to influences like media coverage and testing guidance [10], and differs by population group according to sociodemographic factors [8,9,11–13], geographic location [6,9,14,15], and disease properties [9,12,16]. Crucially, analyses of healthcare-seeking data in combination with sentinel syndromic and influenza case data, have enabled estimates of the true burden of infection and illness [8,17,18] and improvements in forecast accuracy [14]. Similar studies have not (yet) been reported for COVID-19 to our knowledge, despite similar challenges interpreting standard case-based COVID-19 surveillance data.

Here we explore temporal trends in COVID-19 test-seeking behaviour in Australia by symptoms, age group, test type, and geographic location from November 2021–September 2023. We use data from two surveillance systems: a weekly nationwide behavioural survey (NBS hereafter), established by the Australian Government to monitor a range of behavioural responses to COVID-19; and Australia’s FluTracking system, which was adapted in early 2020 to include questions relevant to COVID-19. Our primary analysis focuses on data from the NBS with key metrics then compared to the FluTracking data. In contrast to healthcare-seeking behaviour studies for seasonal influenza or the influenza A (H1N1) 2009 pandemic, our analysis provides insight in the context of mass community testing programs, the widespread use of self-administered rapid antigen tests (RATs; also known as rapid diagnostic tests or lateral flow tests), and significantly greater societal disruption and infection consequences. Finally, the simultaneous deployment of two surveillance systems in Australia — FluTracking and the national survey — enabled comparison of test-seeking behaviour across methodologies.

## Results

### Individuals with symptoms more predictive of COVID-19 were more likely to seek a test

We defined four symptom variables based on their predictability of COVID-19 infection (symptom predictability, see Methods). The symptom variables, from least predictive to most predictive, were defined as individuals reporting: at least one symptom (see Sup Tab.2 for list of symptoms); at least one core symptom (cough, fever, any change in taste or smell, sore throat); at least two core symptoms; and at least fever and cough (which were expected to be the most likely to elicit test-seeking). Respondents of the NBS who reported symptoms more predictive of COVID-19 infection were more likely to seek a test (Fig.1, Sup Tab.4). Over the entire study period (November 2021–September 2023), the proportion of individuals seeking a test was highest in those reporting at least fever and cough, 73.1% (95% Confidence Interval (CI): 71.7%, 74.5%), followed by those reporting at least two core symptoms, 65.0% (95% CI: 64.1%, 65.9%), or at least one core symptom, 51.2% (95% CI: 50.6%, 51.7%), and was lowest in those self-reporting at least one symptom at 37.2% (95% CI: 36.9%, 37.6%). This hierarchy was consistent over the entire study duration, despite changes in test-seeking behaviour over time (Fig.1)

**Figure 1:**
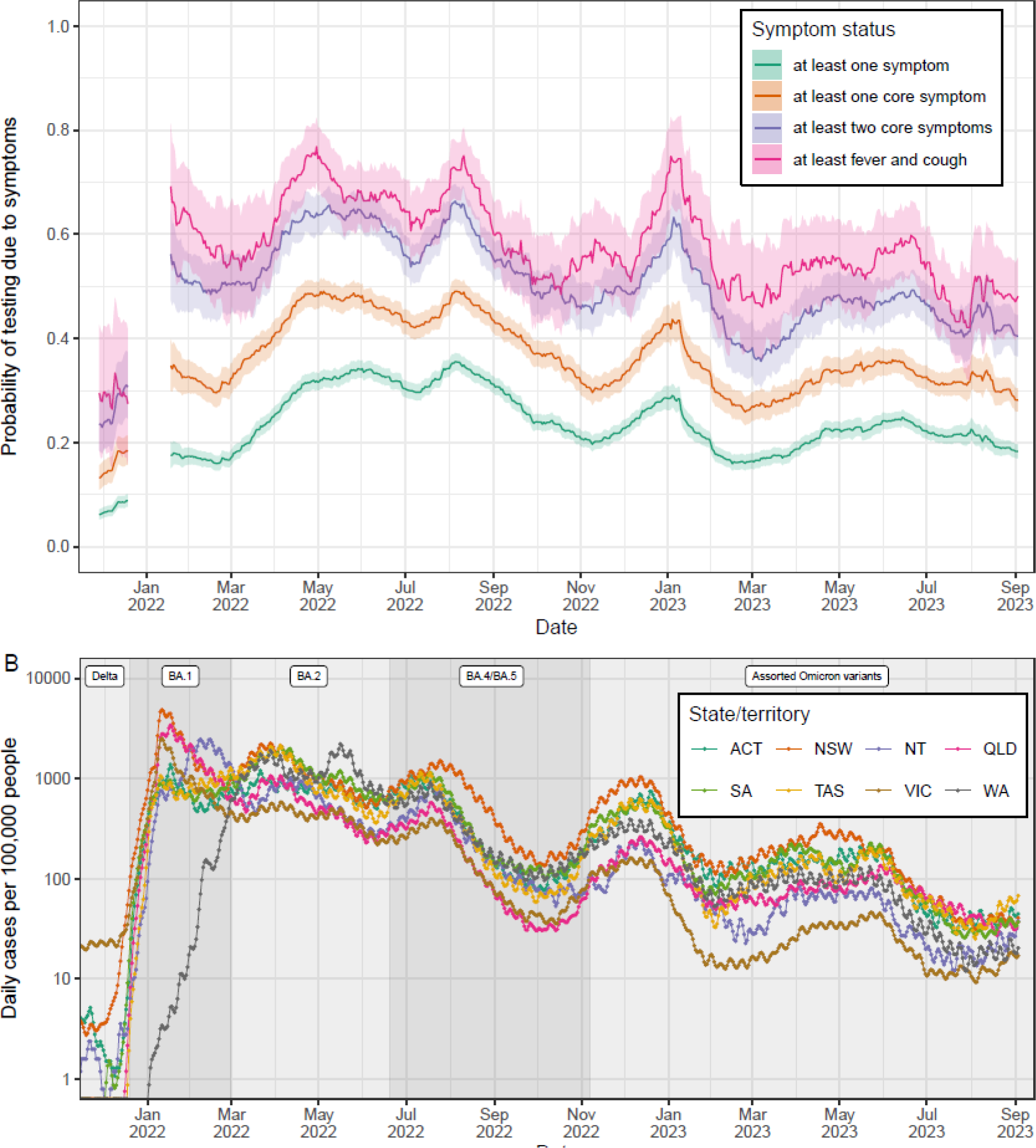
Changes in test-seeking behaviour by symptom status in the NBS. (A) Probability of an individual reporting testing for COVID-19 (with the presence of symptoms given as a reason for testing) given they were exhibiting: at least one symptom (purple); at least one core symptom (orange); at least two core symptoms (pink); or at least fever and cough (green). For each date the proportion of individuals testing due to symptoms for the previous four week period is shown with point estimate (line) and 95% confidence interval (shaded region). Note that no data was collected from 20 December 2021-10 January 2022 inclusive; estimates are not shown from 20 December 2021 to 17 January 2022 (ensures estimates in January 2022 have at least one week of data). (B) The daily number of cases per 100,000 people is shown for the Australian Capital Territory (ACT), New South Wales (NSW), the Northern Territory (NT), Queensland (QLD), South Australia (SA), Tasmania (TAS), Victoria (VIC), and Western Australia (WA). Grey shaded regions show the periods over which each variant (labels) was the dominant variant (greater than 50% of all variants circulating) [36].

For respondents reporting symptoms, the primary reason to seek a test was due to symptoms, irrespective of the symptom predictability (Sup Tab.4). This was consistent over the entire study duration, despite changes in the test-seeking probability over time (Sup Fig.1). For individuals reporting at least one core symptom, the proportion of individuals seeking a test (over the entire study period) due to symptoms was 36.4% (95% CI: 35.8%, 36.9%), followed by contact (close contact with an infected individual), 9.5% (95% CI: 9.2%, 9.9%), occupation (required to take a test as part of their job), 8.3% (95% CI: 8.0%, 8.6%) and ‘other’ (anything other than symptoms, contact or occupation), 4.7% (95% CI: 4.5%, 5.0%). The proportion of individuals who reported seeking a test due to symptoms, contact or their occupation was higher with greater symptom predictability (from at least one symptom to at least fever and cough).

### Coincident peaks in national test-seeking behaviour and case counts

There was a sharp increase in test-seeking behaviour from early December 2021 to late January 2022, coinciding with the escalating phase of the Omicron BA.1 epidemic wave in (most of) Australia (Fig.1). For individuals reporting at least one core symptom the proportion seeking a test due to symptoms increased from 13.2% (95% CI: 10.9%, 15.7%) during 2–29 November 2021 to 34.8% (95% CI: 29.9%, 39.8%) during 11–18 January 2022. There were clear peaks in the proportion seeking a test due to symptoms around April 2022, July 2022, and December 2022, aligning with peaks of epidemic waves driven by the emergence of new Omicron sub-variants. The proportion of individuals seeking a test for reasons other than symptoms (contact, occupation or other) was relatively consistent over time for the duration of the study (Sup Fig.1). However, small increases in the proportion of individuals seeking a test due to contact coincided with peaks in the proportion of individuals seeking a test due to symptoms.

### Heterogeneity in test-seeking behaviour by region

There were clear differences in test-seeking behaviour between individual states and territories in the NBS data (Fig.2B, Sup Tab.5). We considered three distinct time periods within the study period (to have sufficient power to compare individual states/territories): the ‘BA.1/BA.2’ phase (2 November 2021 to 20 June 2022); the ‘BA.4/BA.5’ phase (21 June 2022 to 7 November 2022); and the ‘assorted Omicron variants’ phase (8 November 2022 to 3 September 2023) (Fig.1B). The proportion of individuals (reporting at least one core symptom) seeking a test due to symptoms was lowest in Queensland at 32.6% (95% CI: 30.2%, 35.0%), 37.1% (95% CI: 34.5%, 39.8%) and 26.7% (95% CI: 24.8%, 28.6%) for the three time periods respectively. During the ‘BA.1/BA.2’ phase, the proportion seeking a test due to symptoms was highest in the Australian Capital Territory, 51.1% (95% CI: 46.6%, 55.6%). Whereas in the ‘BA.4/BA.5’ and ‘assorted omicron variants’ phases, the proportion seeking a test due to symptoms was highest in Tasmania, 53.7% (95% CI: 48.2%, 59.1%) and 36.8% (95% CI: 32.7%, 41.0%) respectively.

**Figure 2:**
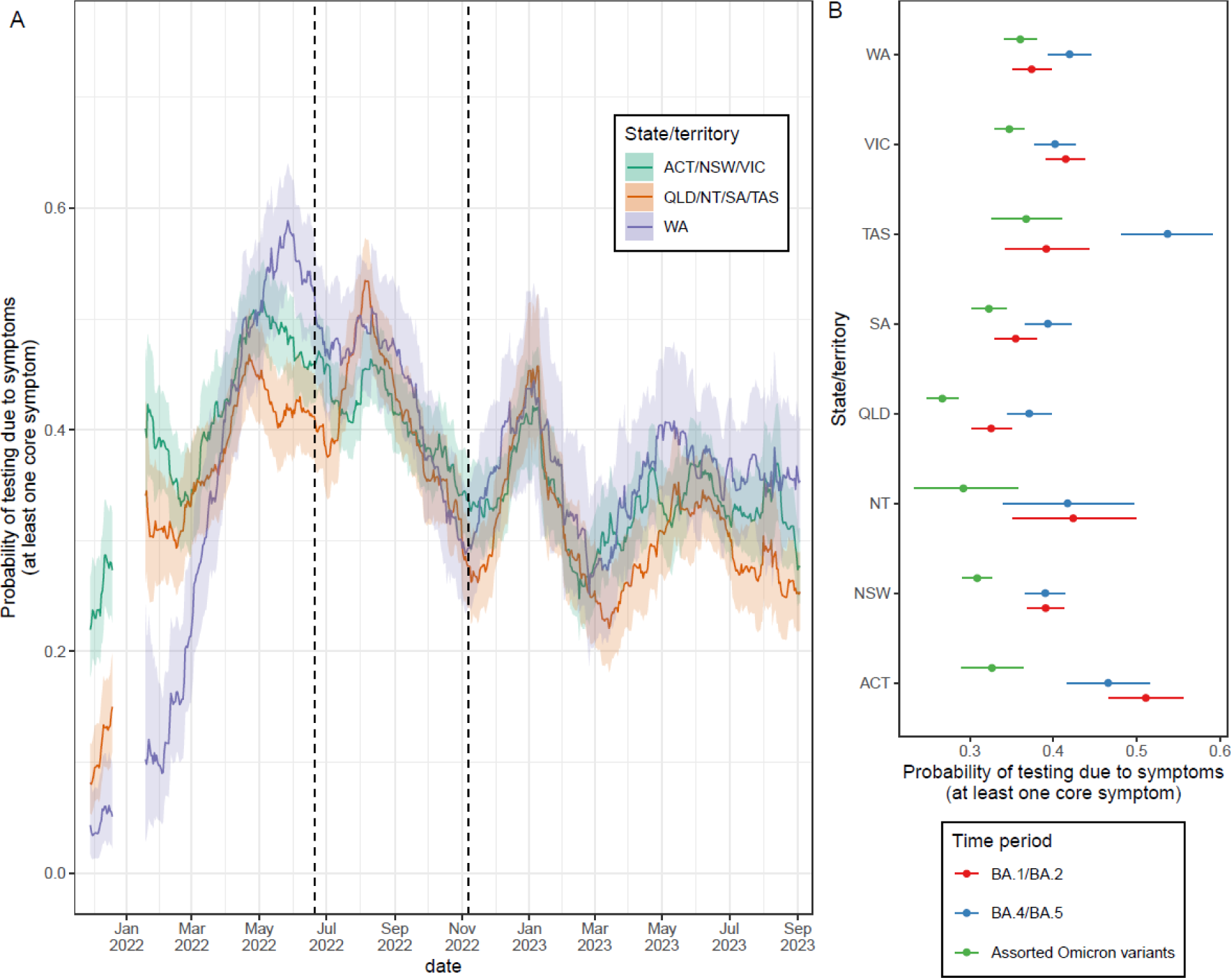
Changes in test-seeking behaviour by state/territory in the NBS. Probability of an individual reporting testing for COVID-19 (with the presence of symptoms given as a reason for testing) given they were exhibiting at least one core symptom. (A) For each date the proportion of individuals testing due to symptoms for the previous four week period is shown with point estimate (line) and 95% confidence interval (shaded region). Note that no data was collected from 20 December 2021-10 January 2022 inclusive; estimates are not shown from 20 December 2021 to 17 January 2022 (ensures estimates in January 2022 have at least one week of data). Estimates are shown for three groupings of states/territories: the Australian Capital Territory (ACT), New South Wales (NSW), and Victoria (VIC); Queensland (QLD), the Northern Territory (NT), South Australia (SA), and Tasmania (TAS); and Western Australia (WA) (B) The average proportion of individuals testing due to symptoms over three periods of the study by state/territory. The three periods chosen represented the BA.1/BA.2 epidemics (2 November 2021 - 20 June 2022), the BA.4/BA.5 epidemic (21 June 2022 - 7 November 2022) and the assorted omicron epidemics (8 November 2022 - 3 September 2023). Point estimates (points) and 95% confidence intervals (lines) are given.

From late-2021 to mid-2022, test-seeking behaviour showed different temporal dynamics between states and territories (Fig.2A). We considered three groupings of states and territories based on differences in epidemic context: New South Wales, Victoria, and the Australian Capital Territory (NSW/VIC/ACT) where widespread community transmission of SARS-CoV-2 (dominated by the Delta variant) had previously established in mid-2021 before the beginning of the study period; Queensland, South Australia, the Northern Territory, and Tasmania (QLD/SA/NT/TAS) where the early phase of the study period represented the first instance of widespread community transmission of SARS-CoV-2 (dominated by the Omicron BA.1 variant) to ever occur in these jurisdictions; and Western Australia (WA) where the first widespread wave of SARS-CoV-2 infections occurred in early-March 2022 (dominated by Omicron BA.2).

The proportion seeking a test due to symptoms during 22 November–19 December 2021 (a period in which Delta was still the dominant variant) was significantly higher in NSW/VIC/ACT, 27.3% (95% CI: 22.6%, 32.5%), compared to QLD/SA/NT/TAS, 15.0% (95% CI: 11.0%, 19.8%), and WA, 5.1% (95% CI: 2.2%, 9.8%). By early-2022, the proportion seeking a test due to symptoms was more comparable between NSW/VIC/ACT and QLD/SA/NT/TAS, both of which were significantly greater than the proportion seeking a test in WA (Fig.2A). In WA, the proportion seeking a test due to symptoms increased greatly during the first half of 2022 reaching a peak of 58.9% (95% CI: 53.6%, 65.0%) from 30 April 2022–27 May 2022. From July 2022 onwards there were similar trends in the proportion seeking a test due to symptoms between states and territories. Temporal trends in the proportion seeking a test due to symptoms in individual states and territories broadly followed similar trends to local case incidence, with similar peak timings (Sup Fig.2).

### Heterogeneity in test-seeking behaviour by age

Test-seeking behaviour of NBS respondents showed clear heterogeneities with age that changed over time (Fig.3B, Sup Tab.6). During the ‘BA.1/BA.2’ phase, the proportion of individuals (reporting at least one core symptom) seeking a test due to symptoms was broadly consistent across age-groups spanning 18–49-year-olds, but lower in older age-groups (50+ years) and lowest in the 65+ age-group at 26.8% (95% CI; 24.1%, 29.6%). Test-seeking behaviour was higher in these older age-groups during the ‘BA.4/BA.5’ phase (compared to the ‘BA.1/BA.2’ phase), and there was greater homogeneity across all age-groups during this phase. Test-seeking behaviour was lower in all age-groups during the ‘assorted omicron variants’ phase (compared to ‘BA.4/BA.5’ phase) with lowest levels in the 18–24 year age-group, 29.0% (95% CI: 27.2%, 30.9%) and highest in the 50-54 year age-group, 37.0% (95% CI: 33.9%, 40.2%). Considering changes over finer temporal timescales (with broader age-groups defined), we found similar temporal trends in the proportion of individuals seeking a test due to symptoms between age-groups (Fig.3A). However, the proportion seeking a test due to symptoms in those aged 60+ was significantly lower than in those aged 18-29 and 30-59 for much of January 2021-July 2021.

**Figure 3:**
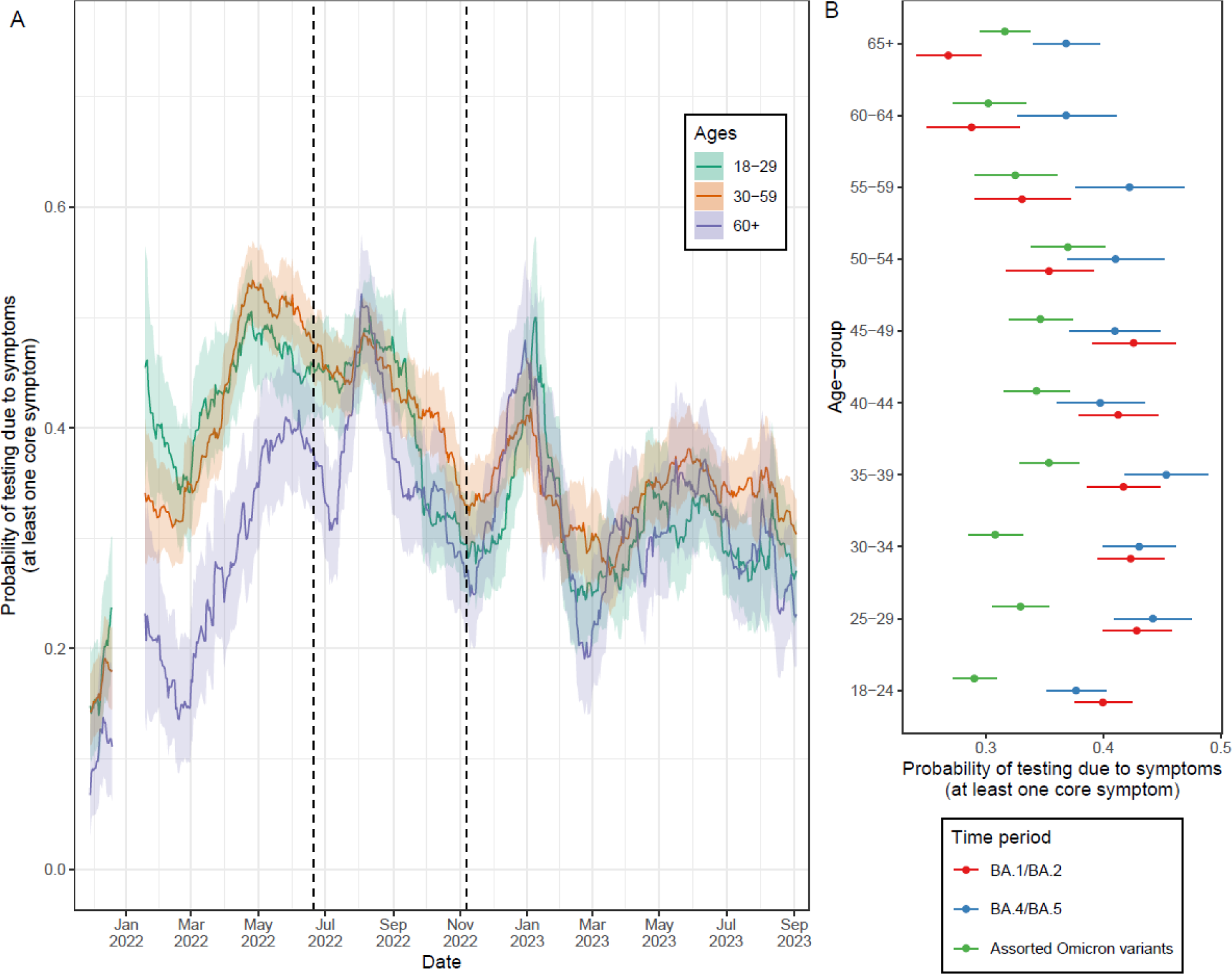
Changes in test-seeking behaviour by age-group in the NBS. Probability of an individual reporting testing for COVID-19 (with the presence of symptoms given as a reason for testing) given they were exhibiting at least one core symptom. (A) For each date the proportion of individuals testing due to symptoms for the previous four week period is shown with point estimate (line) and 95% confidence interval (shaded region). Note that no data was collected from 20 December 2021-10 January 2022 inclusive; estimates are not shown from 20 December 2021 to 17 January 2022 (ensures estimates in January 2022 have at least one week of data). Estimates are shown for three age-groups: 18-29 year olds; 30-59 year olds, 60+ year olds. (B) The average proportion of individuals testing due to symptoms over three periods of the study by finer resolution age-groups. The three periods chosen represented the Delta/BA.1/BA.2 epidemics (2 November 2021 - 20 June 2022), the BA.4/BA.5 epidemic (21 June 2022 - 7 November 2022) and the assorted omicron epidemics (8 November 2022 - 3 September 2023). Point estimates (points) and 95% confidence intervals (lines) are given.

### Propensity to seek a test increased following the introduction of rapid antigen tests

A sharp increase in test-seeking behaviour observed in the NBS from early December 2021 to late January 2022 (Fig.1A) coincided with the wide availability of RATs. During 11–18 January 2022, the proportion of individuals (reporting at least one core symptom) seeking only a PCR test due to symptoms was 15.2% (95% CI: 11.8%, 19.3%), which was comparable to the value of 18.3% (95% CI: 15.7%, 21.3%) during 22 November–19 December 2021 (Fig.4A), when only PCR tests were widely available. However, during 11–18 January 2022, RATs were more widely available in addition to PCR tests, and the proportion seeking only a RAT (due to symptoms), 11.0% (95% CI: 8.0%, 14.6%) or both a PCR and RAT (due to symptoms), 8.6% (95% CI: 5.9%, 11.9%) led to an overall increase in the proportion testing due to symptoms. During January 2022–April 2022, the proportion seeking a RAT due to symptoms increased and the proportion seeking a PCR test due to symptoms decreased to very low levels; this coincided with a time period in which RATs were first introduced at limited capacity and then gradually made more widely available. Over the entire duration of the study for which data on RATs were available (11 January 2022 onwards) the proportion of individuals seeking only a RAT due to symptoms was 26.7% (95% CI: 26.2%, 27.2%). This was significantly higher than the proportion seeking only a PCR test due to symptoms, 4.6% (95% CI: 4.4%, 4.8%) and those seeking both a PCR test and RAT due to symptoms, 5.7% (95% CI: 5.5%, 6.0%).

**Figure 4:**
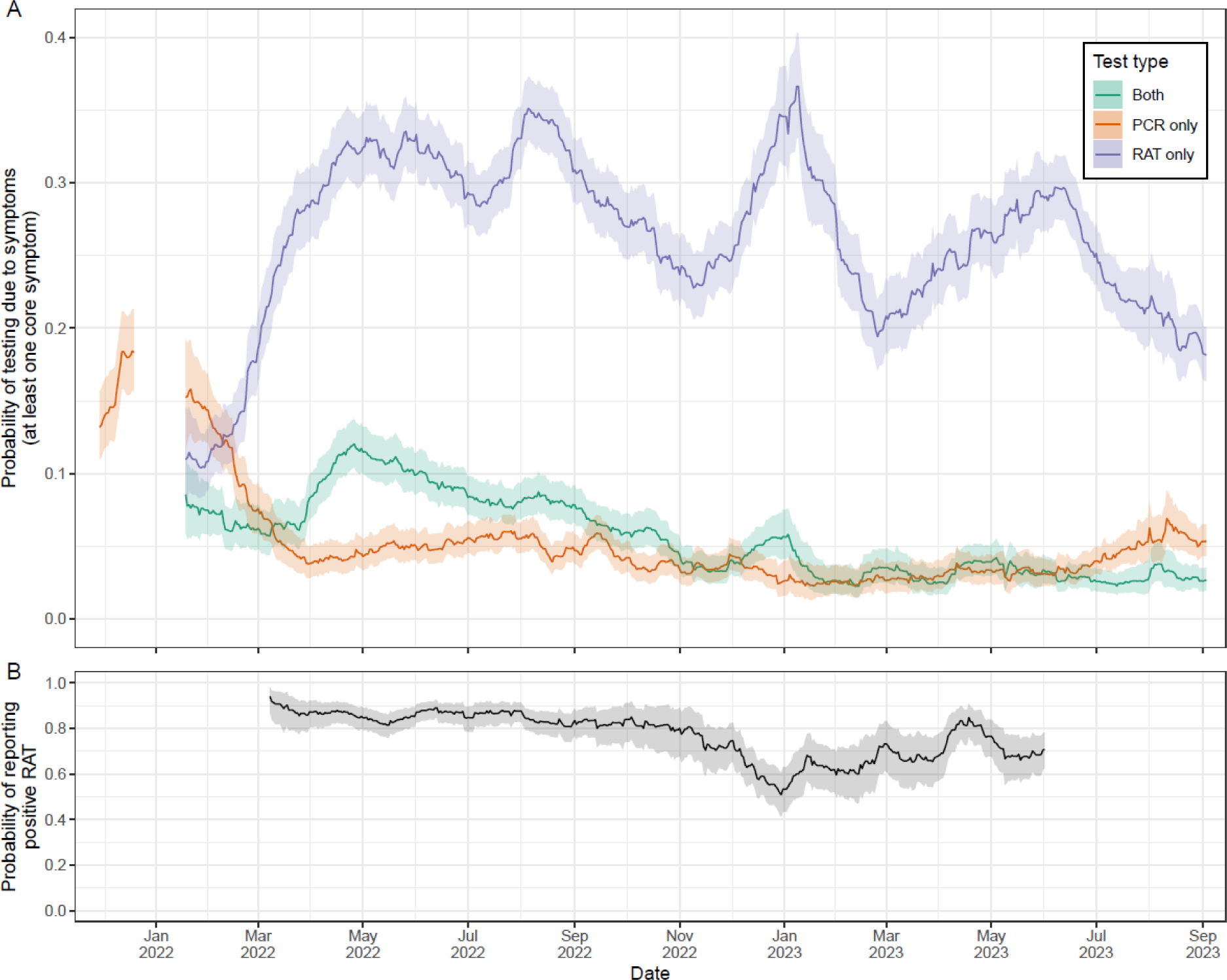
Changes in test-seeking behaviour by test type in the NBS. (A) Probability of an individual reporting testing for COVID-19 (with the presence of symptoms given as a reason for testing) given they were exhibiting at least one core symptom. For each date the proportion of individuals testing due to symptoms for the previous four week period is shown with point estimate (line) and 95% confidence interval (shaded region) by what test they took (PCR test only, RAT only, both RAT and PCR tests). Note that no data was collected from 20 December 2021-10 January 2022 inclusive; estimates are not shown from 20 December 2021 to 17 January 2022 (ensures estimates in January 2022 have at least one week of data). Note that data on Rapid Antigen Testing was not available until 11 January 2022, but there only would have been limited Rapid Antigen Testing in December 2021 as they were not widely available or in use in Australia at that time. (B) The probability of an individual who tested positive for COVID-19 using a RAT reporting their result. For each date the proportion of individuals reporting their positive RAT is shown with point estimates (line) and 95% confidence intervals (shaded region). Note that data on reporting of RAT results was only available from 9 February 2022 and RAT reporting ended in some jurisdictions 1 June 2023; estimates are only shown from 8 March 2022 (four weeks after data first became available) to 1 June 2023.

The probability of an individual (with a positive RAT result) reporting a positive RAT varied over time, though was generally high (Fig.4B; PCR test results did not require reporting by individuals). The probability of reporting a positive RAT was highest during 9 February–8 March 2022 (the first four week period where data were available) at 94.0% (95% CI: 95% CI: 86.7%, 98.0%) and reached a minimum of 50.9% (95% CI: 41.2%, 60.6%) during 3–20 December 2022. On average, over the course of the study for which data were available, the probability of reporting a positive RAT was 79.9% (95% CI: 79.2%, 80.7%).

The probability of a PCR test being positive given symptoms was higher than the probability of a RAT being positive given symptoms (Sup Tab.7). For individuals reporting at least one core symptom, the average probability of a PCR test being positive was 40.3% (95% CI: 38.9%, 41.8%) and the average probability of a RAT being positive was 21.2% (95% CI: 20.5%, 22.0%). The probability of either test being positive was greater for those exhibiting more predictive symptoms of COVID-19 (“at least one symptom” being the least predictive and “at least fever and cough” being the most predictive). The probability of a test being positive given symptoms also varied over time with a large decrease during late 2022 (Sup Fig.4), coinciding with a relative minimum in COVID-19 cases (Fig.1B).

### Differences in test-seeking behaviour across surveillance systems

FluTracking participants were more likely to seek a test due to symptoms than individuals who participated in the NBS over the entire study period. In FluTracking, for individuals self-reporting at least fever and cough, the average probability of seeking a test was 91.8% (91.2%, 92.3%) for those who work with patients and 84.9% (95% CI: 84.6%, 85.2%) for those who do not work face-to-face with patients. These were both higher than the probability of respondents to the NBS (reporting at least fever and cough) seeking a test due to any reason, 73.1% (95% CI: 71.7%, 74.5%), or seeking a test due to symptoms, 58.8% (95% CI: 57.2%, 60.4%). There were similar temporal trends in test-seeking behaviour between the two surveillance systems, with peaks in the proportion of individuals seeking a test broadly aligned (Fig.5).

**Figure 5:**
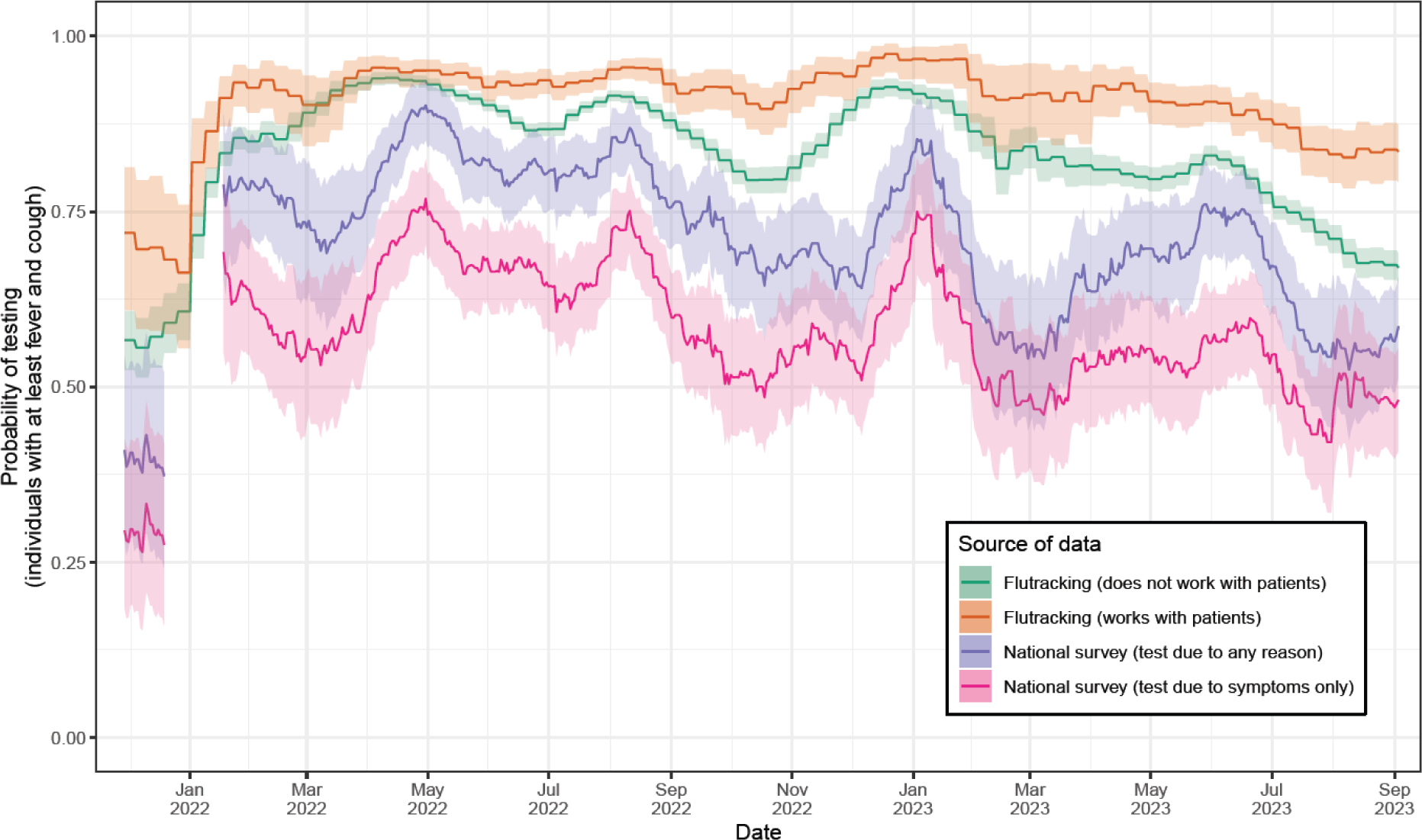
Comparison of the test-seeking behaviour exhibited by participants in the NBS and in the Flutracking surveillance system. Probability of an individual reporting testing for COVID-19 given they were exhibiting at least fever and cough (described as influenza-like illness in Flutracking surveillance system). For each date the proportion of individuals testing due to symptoms for the previous four week period is shown with point estimate (line) and 95% confidence interval (shaded region). Estimates are shown for individuals in the FluTracking study by whether they identified as working face-to-face with patients (orange) or did not identify as working with patients (green). Estimates are shown for individuals taking part in the National survey by whether they reported testing due to symptoms (pink) or reported testing due to any reason (purple). Note that in the National survey no data was collected from 20 December 2021-10 January 2022 inclusive; estimates are not shown from 20 December 2021 to 17 January 2022 (ensures estimates in January 2022 have at least one week of data). Note that FluTracking participants are always asked about the prior week ending Sunday, hence the step-like patterns in the (rolling four-week average) time series.

## Discussion

Using a weekly national behavioural survey (the NBS), we have quantified how test-seeking behaviour changed during the COVID-19 pandemic in Australia from late November 2021–September 2023. Monitoring why individuals seek tests, the rate at which individuals seek and report tests, and how these change over time, is important for informing public health interventions. Changes in test-seeking behaviour will influence the data recorded by case-based surveillance systems. If this is not accounted for, biases may be introduced into analyses of case data, including estimates of the effective reproduction number, forecasts, and scenario projections. Additionally, the effectiveness of some public health interventions will depend on the level of test-seeking behaviour. For example, contact tracing is less effective when test-seeking probabilities are low as the proportion of infections identified (and traceable) will also be low.

Key surveillance indicators generated by case-based surveillance (e.g., the daily number of reported cases) directly depend on test-seeking behaviour. The higher the probability of individuals seeking a test, the higher the number of infections that will be identified as cases—although the exact mathematical relationship can be complex and depends on a myriad of factors (e.g., test sensitivity, symptomatology, and time from infection to test) [19]. Case dynamics are also subject to dynamic behavioural feedback; changes in the number of cases can influence test-seeking behaviour by changing perceptions of disease risk. The increase in test-seeking behaviour that coincided with the start of the Omicron BA.1 epidemic wave in Australia was likely in part due to an increase in perceived (and actual) risk of infection — and of course due to the wider availability of tests (see below). Furthermore, through subsequent epidemic waves, within each jurisdiction we identified transient increases in test-seeking behaviour, with peaks aligning approximately with those in rates of reported cases. Because of these well aligned peaks, it is likely that the size and peakedness of epidemic waves, as measured by case data, were exaggerated relative to if test-seeking behaviour were constant. Further characterisation of the feedback effect between test-seeking behaviour and case rates is warranted but would not be straightforward since the relationship is likely to be highly complex and dynamic through time in response to changes in perceived risk (requiring data on risk perceptions).

Infectious disease dynamics can be highly heterogeneous across populations. Heterogeneity in test-seeking behaviour can obscure or exaggerate the dynamics in case data, potentially leading to poorly targeted public health interventions. We identified differences in test-seeking behaviour by symptom status, jurisdiction, and age-groups. Test-seeking behaviour was higher in respondents reporting more symptoms predictive of COVID-19, which is consistent with previous studies [20]. This means that less symptomatic infections would be underrepresented in data obtained through case-based surveillance. Additionally, any changes to the symptomatology of infections through time will influence case data. During the COVID-19 pandemic, the accumulation of vaccine- and infection-acquired immunity has reduced the average number and duration of symptoms exhibited by infected individuals [21]. Different variants have also shown differences in overall symptom rates [22], and in the rate of “core” COVID-19 symptoms [23,24].

In our study, the probability of seeking a test was lower in younger and older individuals (only adults considered). Where infection-prevalence surveys were conducted throughout the COVID-19 pandemic (the United Kingdom), infection rates were often higher in younger individuals [25], and the severity of infection was greater in older individuals [4]. In combination with age-specific variability in test-seeking behaviour, infection rates in younger individuals could have been under-represented in case data by lower rates of testing, while estimates of severity of infection in older individuals generated from case data could have been overestimated (i.e., lower number of cases detected, but deaths/hospitalisations detected consistently). Differences in test-seeking behaviour were also observed between Australian states and territories, which may have generated a misleading picture of relative levels of epidemic activity across jurisdictions. For example, differences in case rates between jurisdictions in late-2021/early-2022 may have been exaggerated or masked by the relatively greater differences in test-seeking behaviour during this period.

There are obvious public health benefits from high levels of test-seeking behaviour. The higher the probability of testing given symptoms, the higher the probability of detecting infected individuals (greater case ascertainment), and in turn greater reductions in transmission can be achieved through case isolation and tracing and quarantining of close contacts. Additionally, a greater proportion of infected individuals can gain access to therapeutics and, if contact tracing is sufficiently fast,earlier in the course of infection, reducing the number of severe infections. Thus, changes in test-seeking behaviour can alter the underlying epidemic dynamics and disease outcomes, as well as the data-generating processes by which these epidemic dynamics are measured (e.g., case-based surveillance).

The rapid increase in test-seeking behaviour observed across Australia in early-2022, which continued to slowly climb until mid-2022, coincided with wider availability of RATs. This increased availability was positive for public health; more infections could be identified, and individuals could better inform their own actions (e.g., could test before social events, could choose to self-isolate if positive, access treatment if positive, etc). However, the use of RATs also introduced additional layers of complexity for assessing how behaviour impacted case numbers. Firstly, whereas data on PCR test results is collected and reported to health authorities by laboratories, RATs are performed at home and require self-reporting of positive results. Unsurprisingly, we found that not all individuals report their test results; the probability of an individual reporting a positive test was about 80% nationally and varied over time. Secondly, RATs are less sensitive than PCR tests [26] and so although more individuals were testing overall, the probability of an infected individual (who sought any test) testing positive would have declined. We found that the probability of testing positive given symptoms was lower for RATs relative to PCR tests which supports this difference in sensitivity. Overall, the wide use of two types of tests, with different test-seeking behavioural patterns and sensitivities, introduces an additional layer of complexity when analysing trends in case data.

Our study has limitations. The NBS only surveyed adults and was administered online and in English, limiting the individuals who were able to take part, reducing the study’s representativeness of the general population. Additionally, the relative sampling rates of states and territories, meant that some jurisdictions were under- and over-represented (note that this would not have affected comparisons within and between states and territories). Similar behavioural studies have also been limited in their representativeness, with women and older individuals—who may be more health conscious (see below)—often being heavily over-represented [27,28]. However, the NBS was far more representative in terms of age (post-18 years old) and gender compared to similar studies. In particular, the Male to Female ratio of participants (a ratio of 1 representing equal numbers) in the NBS was 0.92, compared to 0.68 for FluTracking, and 0.48 (for participants identified with symptoms) for the ZOE study in England [20].

The NBS had fixed weekly proportional sampling quotas based on key demographic variables. In contrast, many behavioural studies seek to recruit as many participants as possible [28] but give less consideration to ensuring that participants are representative of the general population. It has been suggested that participatory surveillance systems typically include more health-conscious individuals [20] since the cohort are specifically recruited to be part of a health-related survey. The recruitment process of the NBS may have resulted in a more representative subset of the population in terms of health consciousness; individuals were recruited to participate in surveys of public opinion on a wide range of topics and were remunerated for their participation. We identified differences in test-seeking behaviour between individuals in FluTracking and the NBS which may be reflective of sociodemographic differences in the study populations. There was a greater probability of seeking a test for individuals in FluTracking — this was even more pronounced for those who worked directly with patients (e.g., in healthcare). This highlights the importance of studies in which data are collected through multiple methods. Mixed methods designs would allow estimates between methodologies to be compared, strengthening findings where results are similar, and highlighting uncertainty or potential biases where differences are observed. For example, social desirability bias — the tendency for survey participants to self-report in line with social norms and expectations rather than their true opinions or behaviours (e.g., potentially biassing responses in the NBS towards ’yes I reported my positive RAT’) — is known to vary by survey mode (e.g., anonymous versus face-to-face interviews) [29].

Monitoring test-seeking behaviour is important for understanding the data generating processes of case-based surveillance systems, and for directly informing public health interventions. Since early-2022, analyses of data collected by the NBS have been reported weekly to Australia’s key national decision-making bodies [30]. Our study has demonstrated that during the COVID-19 pandemic there was substantial heterogeneity in test-seeking behaviour in Australia between age-groups, jurisdictions, and type of test, and through time. Continued collection of behavioural data through dedicated surveillance systems, where careful consideration is given to achieving representativeness of the target population, is important for understanding the future transmission dynamics of SARS-CoV-2.

## Methods

### Epidemiological and policy context

The study period, late November 2021 to September 2023, spans several COVID-19 epidemic phases and policy transitions in Australia, with variability across Australia’s eight states and territories.

In July 2021, transmission of the Delta variant became established in the state of New South Wales (NSW), followed by Victoria (VIC) and the Australian Capital Territory (ACT), resulting in widespread waves of infection in these jurisdictions. By late November 2021, the beginning of our study period, the initial waves of Delta infection had peaked and were in decline. In late November 2021, the Omicron variant emerged in Australia. This occurred during a period in which jurisdictions were transitioning from a response objective of minimising community transmission, largely through strict public health and social measures, to one of manageable transmission in terms of health system capacity (the “re-opening transition”) [2,31]. The timing of the re-opening transition varied across Australian states and territories, resulting in notable differences in COVID-19 epidemiology during the earliest phase (late November 2021–May 2022) of our study period. Nonetheless, community transmission was widespread in all jurisdictions by early-March 2022 (Figure 1B).

As a result of the re-opening transition, most border and social restrictions had been lifted by the beginning of our study period, two dose vaccination coverage was greater than 85% nationally (in ages 16 years and above), and test-trace-isolate-quarantine strategies were in place [32], supported by mass community testing programs. Specific population groups, such as those with respiratory symptoms and contacts of confirmed COVID-19 cases, were encouraged to seek testing [33], although any individual could access free polymerase chain reaction (PCR) for COVID-19 from one of many geographically distributed testing clinics, without consulting a health practitioner.

The rapid growth of the Omicron BA.1 epidemic across most of the country in December 2021, and the reopening of many interstate borders with a requirement for a negative PCR test resulted in testing demand that quickly outstripped PCR testing and contact tracing capacity, leading to testing bottlenecks and delays in both test access and processing time. This limited the usefulness of mass PCR testing motivated an urgent need for a supplementary pathway to testing. Rapid antigen tests (RATs; also known as rapid diagnostic tests or lateral flow tests) had been available in Australia since late 2021, but only became widely available in mid-January 2022, alleviating the pressure on PCR testing clinics. Online portals for self-reporting positive RATs were also established by jurisdictional authorities at around the same time.

Throughout 2022, remaining border and social restrictions were gradually lifted, and test-trace-isolate-quarantine policies were scaled back. Isolation mandates for confirmed cases ended nationally on 14 October 2022, and by early May 2023, all Australian jurisdictions had discontinued referral-free community PCR testing for COVID-19, prioritising PCR testing for those most vulnerable to severe illness and death (via request from a health practitioner). RATs remained available but reporting of positive results was no longer mandatory and online reporting portals were gradually stood down in most jurisdictions during the second half of 2023.

### National Behaviour Survey data

The National Behaviour Survey (NBS) is a weekly cross-sectional online survey of behaviours relating to COVID-19 risks and public health measures. The survey was first fielded in early April 2020 with 1,000 participants per week. The sample size was increased to 2,000 participants per week in mid-May 2020. The questionnaire is administered by a commercial market research company (Painted Dog Research) on behalf of the Australian Government Department of Health and Aged Care. Survey participants are sampled from a panel of over 160,000 Australian adults who have agreed to participate in online surveys of public opinion, recruited by market research companies Dynata and Octopus. The survey is only available to participants in English. Proportional sampling quotas are used based on Australian Bureau of Statistics census data [34] to ensure that respondents represented the Australian adult population based on age, gender, and location (state and metropolitan or regional). Weekly quotas are outlined in Supplementary Table 1.

The survey includes a range of questions on behaviours relevant to COVID-19 transmission, surveillance, and control. Some of the survey items remained consistent for each wave of the survey, others were added, dropped, or adjusted to ensure relevance to public health issues at the time. Our analysis uses responses to five questions around whether participants experienced COVID-19 symptoms, sought a test, reported a positive RAT result, and their reason for testing (a copy of the full questions and responses options are included in Supplementary Table 2).

Individual-level data including responses to the questions, basic demographic information (respondent’s state/territory of residence, age, and gender) and the date on which the survey was completed were obtained from 2 November 2021 to 3 September 2023. The overall number of participants by state/territory, age-group and gender are provided in Supplementary Table 3.

### Flutracking data

FluTracking is a participatory surveillance system that contributes to community-level influenza and COVID-19 surveillance in Australia and New Zealand through weekly online voluntary surveys. Participants are recruited online through workplace campaigns, existing participants, the FluTracking website, traditional media, and social media promotion. After registering as a “FluTracker”, participants receive a weekly email with a link to an online questionnaire. The questionnaire includes questions on the presence of typical influenza and COVID-19 symptoms during the past week, whether they sought a test and/or medical attention, and test results (see https://www.flutracking.net/Demo/AU for demonstration). Over 150,000 Australian and New Zealand participants completed at least one survey in 2022.

Weekly counts of completed FluTracking surveys, self-reported presence of influenza-like illness (fever and cough, ILI hereafter) in the most recent reporting week, and self-reported COVID-19 tests obtained from the week ending 7 November 2021 to the week ending 3 September 2023. Note that there is an option for participants to take a break from completing surveys over October-April each year; the sample size will be reduced over this period. The overall number of survey records over the study period by state/territory, age-group and sex are provided, and compared to the actual distributions based on Australian census data [35] in Supplementary Table 3.

### Statistical analyses

Individuals in the NBS were split into binary symptom categories based on them reporting specific symptoms or not. Four different symptom variables were defined based on their predictability of COVID-19 infection (symptom predictability). The symptom variables, from least predictive to most predictive, were defined as individuals reporting: at least one symptom (see Sup Tab.2 for list of symptoms); at least one core symptom (cough, fever, any change in taste or smell, sore throat); at least two core symptoms; and at least fever and cough. The four core symptoms were selected because they were highly predictive of SARS-CoV-2 infection across all variants circulating during the study period, although the most predictive symptom varied by SARS-CoV-2 variant [22]. Any change in taste or smell was the most predictive symptom for the Delta variant. Sore throat was more predictive of BA.2 infection than all other symptoms (excluding fever and cough). Finally, fever and cough were the most predictive symptoms of BA.1 and BA.2 infection. We expected these core symptoms—particularly fever and cough— to elicit greater test-seeking behaviour as public health messaging was often targeted at symptoms perceived to be more predictive of SARS-CoV-2 infection. An individual could be in multiple of these symptom categories (e.g., an individual reporting a fever and cough would be in all categories).

The probability of an individual seeking a test given they were in a specific symptom category was estimated by calculating the proportion of individuals in the symptom category who reported seeking a test. Similarly, the probability of individuals (in a specific symptom category) seeking a test due to specific reasons (symptom, contact, job and other) were also estimated. Exact 95% confidence intervals were calculated using the Clopper-Pearson method.

The probabilities of individuals (self-reporting at least one core symptom) seeking a test due to symptoms were estimated for each state/territory, for different age-groups, and for the type of test sought (only a RAT, only a PCR test, both RAT and PCR test).

For individuals (within a specific symptom category) who reported taking either a PCR test or RAT, the probability of the respective test being positive was estimated. For all individuals who had a positive RAT, the probability of the positive RAT being self-reported via an online portal was also estimated (PCR test results were systematically reported by laboratories).

The probability of individuals in the FluTracking surveillance system who self-reported ILI (fever and cough) seeking a test was estimated for the subset of participants who work/do not work with patients (note that this question is only asked for participants aged 15 years or older, those aged under 15 years are assumed to not work with patients). We compared these estimates to estimates of the probability of participants (self-reporting fever and cough) seeking a test due to any reason or seeking a test due to their symptoms, in the NBS.

Trends over time for all probabilities were estimated by calculating the proportion and 95% confidence intervals (as above) over a rolling four-week time window. Note that the NBS did not collect any data for three weeks from 20 December 2021 to 10 January 2022 and so estimates are not presented over this period and are not presented until at least 18 January so that the four-week time window includes at least a week of data. For questions in which data only began being collected midway through the study period, estimates of probabilities are only presented for a period in which there were four weeks of data available.

Analysis was performed in the R statistical computing environment (version 4.2.3) [37]. Binomial confidence intervals were calculated using the epitools package (version 0.5.10.1). [38] Figures were created using the ggplot2 package (version 3.4.2) [39].

## Data availability

Access to the individual-level survey data from the NBS and FluTracking is restricted to protect participants’ anonymity. The aggregated values and confidence intervals for all figures and supplementary figures are provided in the supplementary materials.

## Code availability

Code that performed the analyses and generated the figures is available at https://github.com/Eales96/test-seeking-behaviour/

## Ethics statement

This study was approved by the University of Melbourne Human Research Ethics Committee (reference number 2023-26949-40340-2)

## Acknowledgements

The authors wish to acknowledge participants of the NBS and FluTracking systems.

Our analyses use surveillance data reported through the Communicable Diseases Network Australia (CDNA) as part of the nationally coordinated response to COVID-19. We thank public health staff from incident emergency operations centres in state and territory health departments, and the Australian Government Department of Health and Aged Care, along with state and territory public health laboratories. We thank members of CDNA for their feedback and perspectives on the results of the analyses. Funding for this work was provided by the Australian Government Department of Health and Aged Care and the National Health and Medical Research Council of Australia through the Investigator Grant Scheme (FMS Emerging Leader Fellowship, 2021/GNT2010051)

## Figures

**Supplementary Figure 1:**
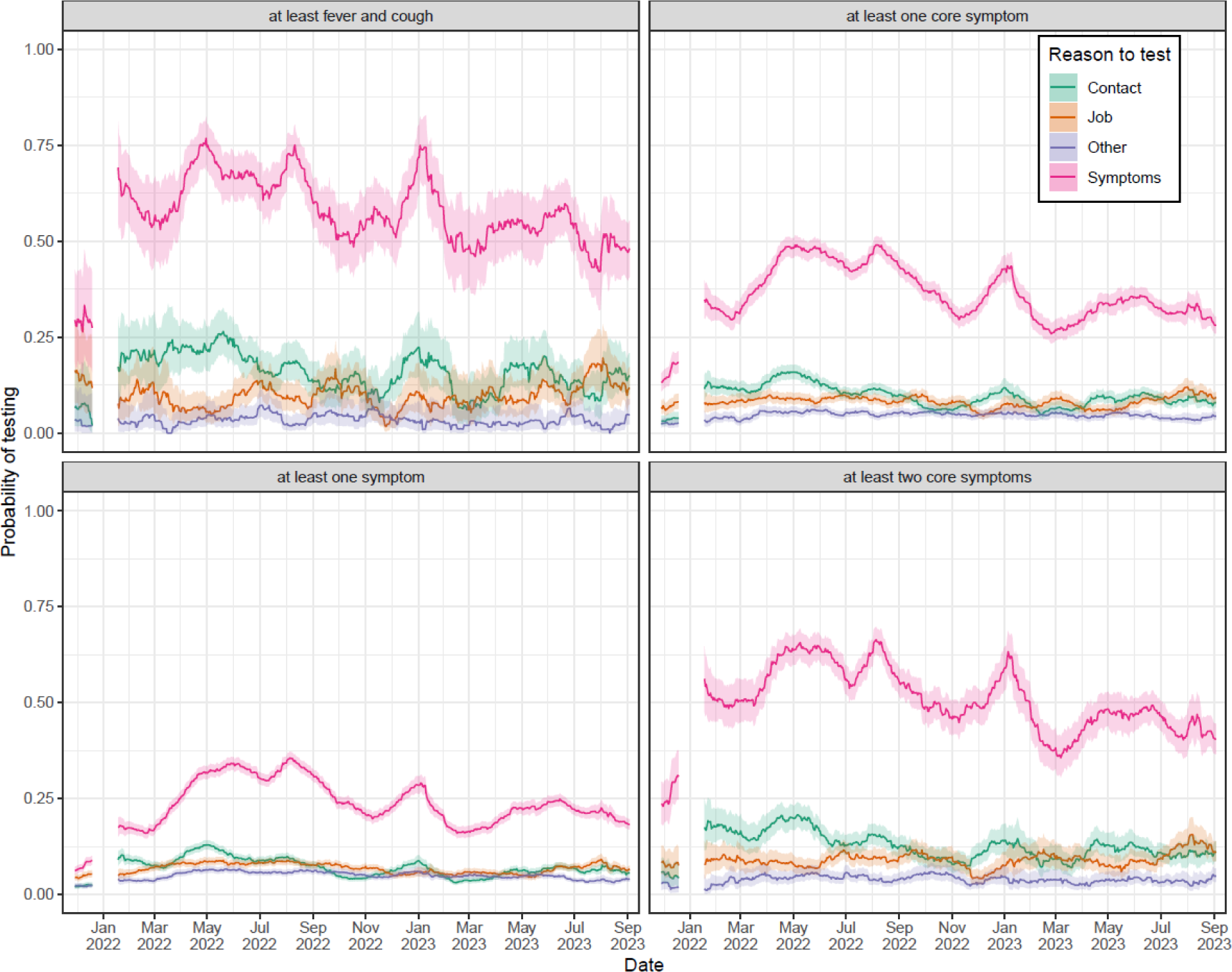
Changes in test-seeking behaviour by reason to test. Probability of an individual reporting testing for COVID-19 due to: the presence of symptoms (pink); contact with an infected individual (green); testing being a requirement of their job (orange); and any other reason (purple). Estimates are shown for individuals that were exhibiting: at least one symptom (bottom-left panel); at least one core symptom (top-right panel); at least two core symptoms (bottom-right panel); or at least fever and cough (top-left panel). For each date the proportion of individuals testing (due to the specific reason) for the previous four week period is shown with point estimate (line) and 95% confidence interval (shaded region). Note that no data was collected from 20 December 2021-10 January 2022 inclusive; estimates are not shown from 20 December 2021 to 17 January 2022 (ensures estimates in January 2022 have at least one week of data).

**Supplementary Figure 2:**
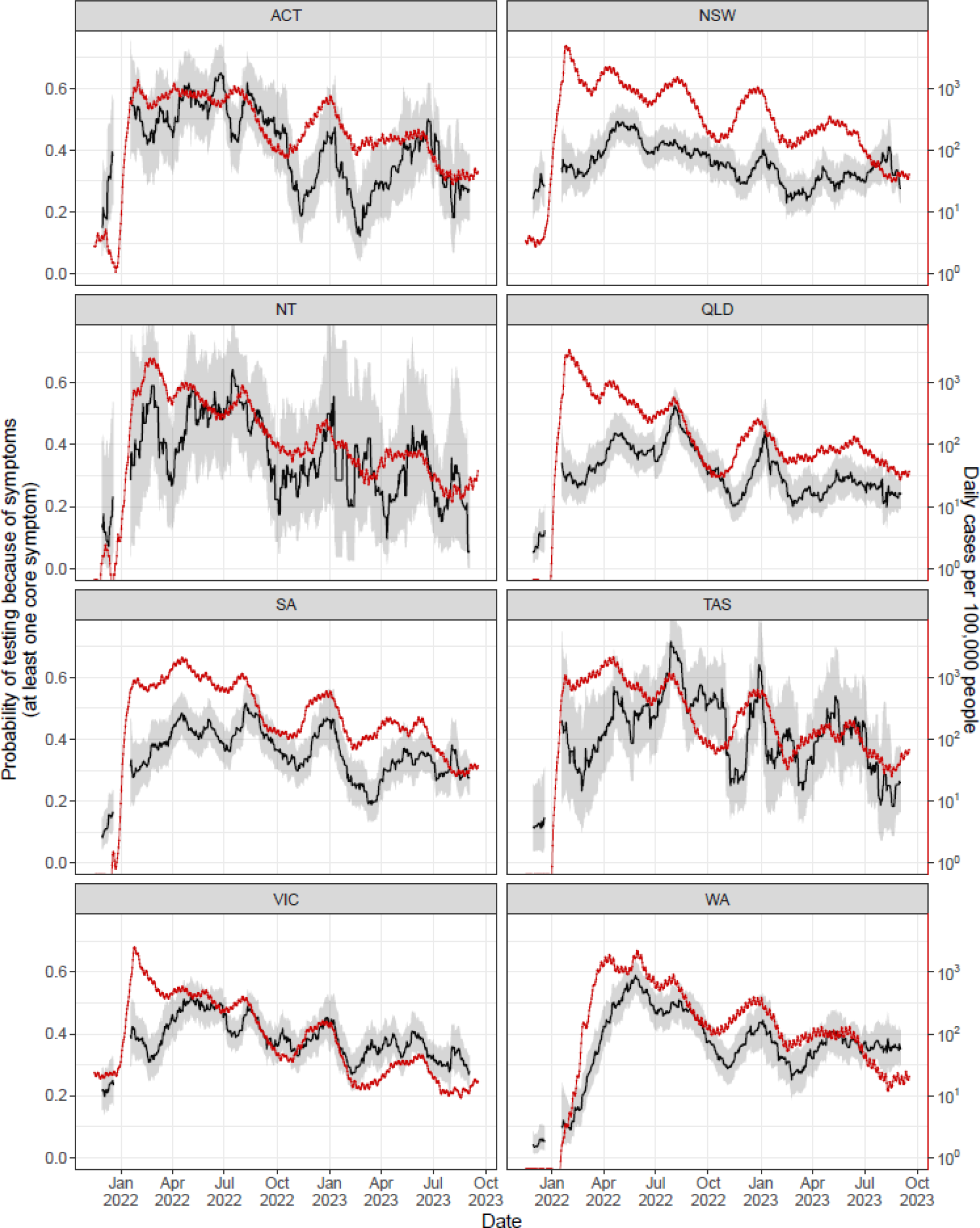
Changes in test-seeking behaviour and case rates over time by state. Probability of an individual reporting testing for COVID-19 (with the presence of symptoms given as a reason for testing) given they were exhibiting at least one core symptom (left-hand axis). For each date the proportion of individuals testing due to symptoms for the previous four week period is shown with point estimate (black line, left-hand axis) and 95% confidence interval (shaded region, left-hand axis). Note that no data was collected from 20 December 2021-10 January 2022 inclusive; estimates are not shown from 20 December 2021 to 17 January 2022 (ensures estimates in January 2022 have at least one week of data). Also shown is the daily number of cases per 100,000 people (red points, red line, right-hand axis). The time series for the daily number of cases per 100,000 people has been translated by two weeks along the x-axis (to the right) to better match the dates for the probability of testing due to symptoms (rolling previous four-week average). Estimates are shown for the Australian Capital Territory (ACT), New South Wales (NSW), the Northern Territory (NT), Queensland (QLD), South Australia (SA), Tasmania (TAS), Victoria (VIC), and Western Australia (WA).

**Supplementary Figure 3:**
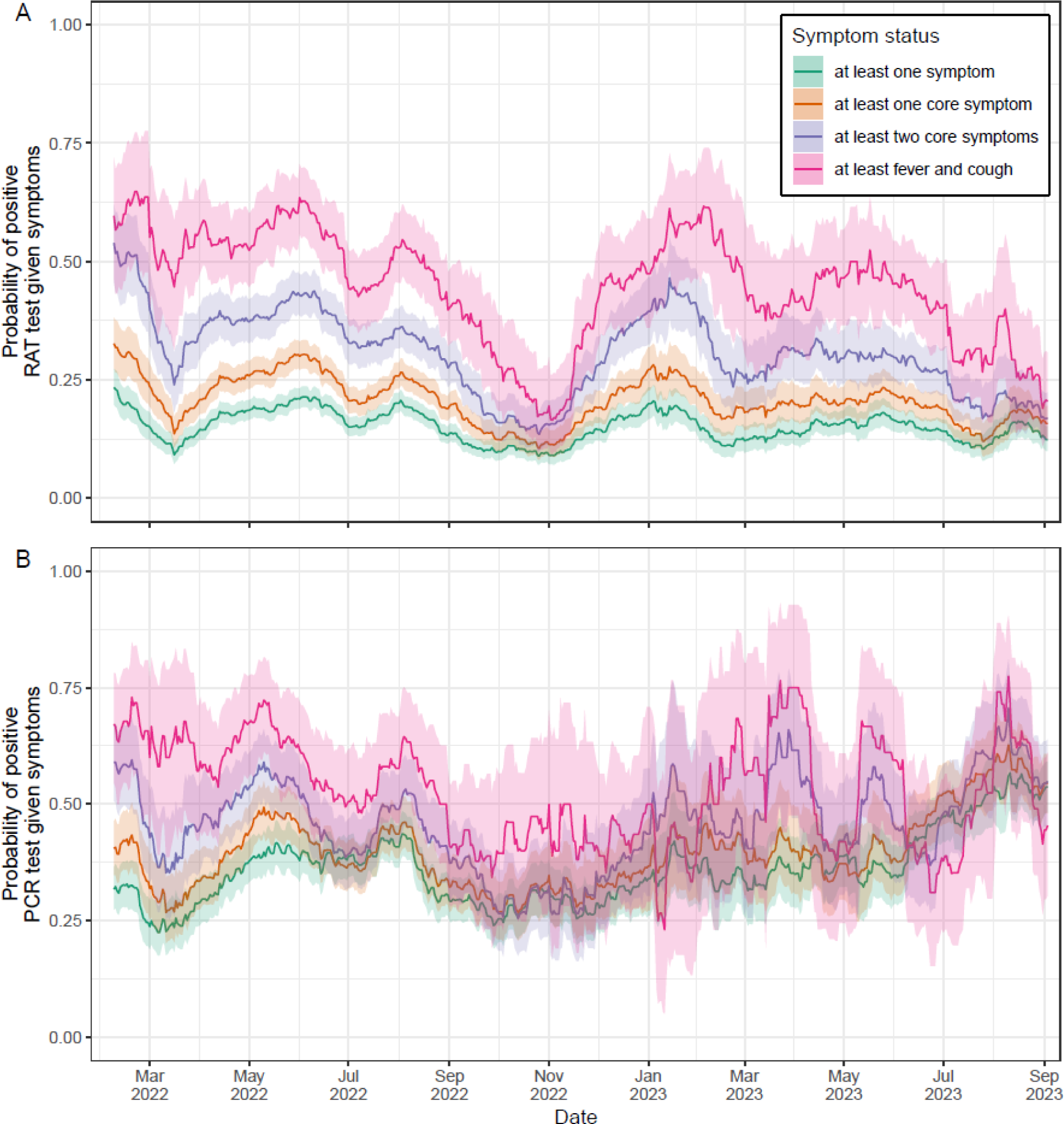
Changes in the probability of a test being positive for symptomatic individuals over time. Probability of a (A) RAT and (B) PCR test returning positive results given the individuals testing are exhibiting: at least one symptom (purple); at least one core symptom (orange); at least two core symptoms (pink); or at least fever and cough (green). For each date the proportion of tests that are positive for COVID-19 for the previous four week period is shown with point estimate (line) and 95% confidence interval (shaded region). Note that data was only available on test positivity from 11 January 2022 and estimates are only shown from 7 February 2022 (four weeks after data first became available).

**Supplementary Table 1:** Weekly study recruitment quotas throughout the analysis period.

| Region | N | Age/Gender | Male | Female | Total | City | N |
| --- | --- | --- | --- | --- | --- | --- | --- |
| NSW | 400 | 18-24 | 120 | 116 | 236 | Major city | 1560 |
| VIC | 400 | 25-34 | 182 | 186 | 368 | Regional | 440 |
| QLD | 350 | 35-44 | 170 | 176 | 346 | Total | 2000 |
| WA | 350 | 45-54 | 166 | 172 | 338 |  |  |
| SA | 350 | 55-64 | 147 | 156 | 303 |  |  |
| ACT | 90 | 65+ | 190 | 219 | 409 |  |  |
| TAS | 80 | Total | 975 | 1025 | 2000 |  |  |
| NT | 30 |  |  |  |  |  |  |
| Total | 2000 |  |  |  |  |  |  |

**Supplementary Table 2:** Survey questions used in the analysis. Core symptoms in the responses to the first question are marked with an asterisk.

| Question | Response options |
| --- | --- |
| Have you experienced any of the following symptoms during the past two weeks?<br>*Select all that apply.* | ☐ Cough*<br>☐ Fever*<br>☐ Difficulty breathing<br>☐ Sore throat*<br>☐ Tiredness<br>☐ Joint aches<br>☐ Headache<br>☐ Runny or stuffy nose<br>☐ Any change in taste or smell*<br>☐ Nausea and/or vomiting<br>☐ Chills<br>☐ I have not experienced any of these symptoms in the past two weeks |
| Did you have a PCR test (e.g., at a drive-through, respiratory clinic or hospital that is sent to a lab) or a Rapid Antigen Test (RAT) (e.g., at home, work, school, etc)?<br>*Select all that apply.* | ☐ PCR test<br>☐ Rapid Antigen Test (RAT)<br>☐ I don't know. |
| What was the result of your most recent PCR test? *Select the option that best applies.*<br>This question is only asked of respondents who report seeking a PCR. An equivalent question is asked of respondents who report seeking an antigen test. | ☐ The test showed I have COVID-19 (positive result)<br>☐ The test showed I do not have COVID-19 (negative result)<br>☐ The test result was inconclusive<br>☐ I'm still waiting to hear the result<br>☐ Prefer not to say |
| Why did you get a COVID-19 test? *Select all that apply.*<br>This question is only asked of respondents who report seeking either a PCR or antigen test or both. | ☐ I experienced symptoms consistent with COVID-19 infection<br>☐ I was informed I had been in contact with a confirmed COVID-19 case<br>☐ It was required as part of my job or occupation<br>☐ Other reason (please specify) |
| Did you report your positive rapid antigen test (RAT) result to a state/territory health department?<br>This question is only asked of respondents who report a positive RAT result. | ☐ Yes<br>☐ No |

**Supplementary Table 3:**
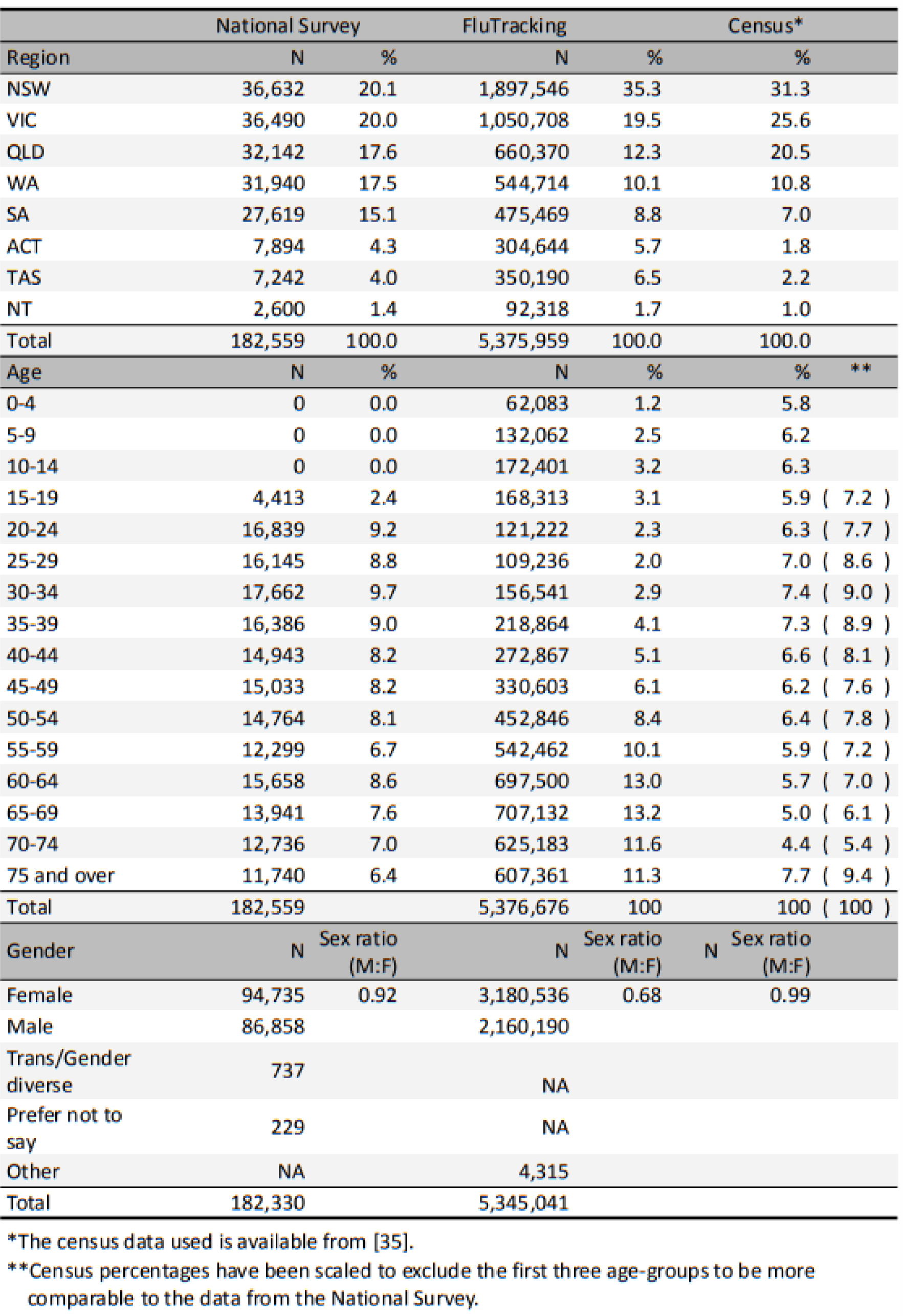
Number of participants by state/territory, age-group and gender for the National Survey and FluTracking study.

**Supplementary Table 4:** The probability of testing by reason to test. The average (over entire study duration) probability of an individual (by symptom status) reporting testing for COVID-19 due to: any reason (any); the presence of symptoms (symptom); contact with an infected individual (contact); testing being a requirement of their job (job); and any other reason (other).

| Symptoms | Reason to test | Proportion (95% confidence interval) |
| --- | --- | --- |
| At least one symptom | Any | 0.372 ( 0.369 , 0.376 ) |
|  | Contact | 0.069 ( 0.067 , 0.071 ) |
|  | Other | 0.049 ( 0.047 , 0.050 ) |
|  | Symptoms | 0.234 ( 0.231 , 0.237 ) |
|  | Job | 0.068 ( 0.066 , 0.070 ) |
| At least one core symptom | Any | 0.512 ( 0.506 , 0.517 ) |
|  | Contact | 0.095 ( 0.092 , 0.099 ) |
|  | Other | 0.047 ( 0.045 , 0.050 ) |
|  | Symptoms | 0.364 ( 0.358 , 0.369 ) |
|  | Job | 0.083 ( 0.080 , 0.086 ) |
| At least two core symptoms | Any | 0.650 ( 0.641 , 0.659 ) |
|  | Contact | 0.127 ( 0.121 , 0.133 ) |
|  | Other | 0.040 ( 0.037 , 0.044 ) |
|  | Symptoms | 0.512 ( 0.503 , 0.521 ) |
|  | Job | 0.091 ( 0.085 , 0.096 ) |
| At least fever and cough | Any | 0.731 ( 0.717 , 0.745 ) |
|  | Contact | 0.162 ( 0.150 , 0.174 ) |
|  | Other | 0.036 ( 0.030 , 0.042 ) |
|  | Symptoms | 0.588 ( 0.572 , 0.604 ) |
|  | Job | 0.097 ( 0.088 , 0.107 ) |

**Supplementary Table 5:** The probability of testing due to symptoms by state/territory. The average (over three separate periods of the study) probability of an individual (by state/territory) reporting testing for COVID-19 (with the presence of symptoms given as a reason for testing) given they were exhibiting at least one core symptom.

| State/territory | Proportion (95% confidence interval) |  |  |
| --- | --- | --- | --- |
|  | 2021-11-02 to 2022-06-20 | 2022-06-21 to 2022-11-07 | 2022-11-08 to 2023-09-03 |
| NSW | 0.391 ( 0.369 , 0.413 ) | 0.390 ( 0.367 , 0.415 ) | 0.309 ( 0.292 , 0.327 ) |
| VIC | 0.415 ( 0.392 , 0.438 ) | 0.402 ( 0.378 , 0.427 ) | 0.347 ( 0.330 , 0.365 ) |
| ACT | 0.511 ( 0.466 , 0.556 ) | 0.466 ( 0.417 , 0.515 ) | 0.327 ( 0.290 , 0.364 ) |
| QLD | 0.326 ( 0.302 , 0.350 ) | 0.371 ( 0.345 , 0.398 ) | 0.267 ( 0.248 , 0.286 ) |
| SA | 0.355 ( 0.330 , 0.380 ) | 0.394 ( 0.366 , 0.422 ) | 0.323 ( 0.303 , 0.344 ) |
| NT | 0.424 ( 0.352 , 0.499 ) | 0.417 ( 0.341 , 0.497 ) | 0.292 ( 0.233 , 0.357 ) |
| TAS | 0.392 ( 0.342 , 0.443 ) | 0.537 ( 0.482 , 0.591 ) | 0.368 ( 0.327 , 0.410 ) |
| WA | 0.374 ( 0.351 , 0.398 ) | 0.420 ( 0.394 , 0.445 ) | 0.361 ( 0.341 , 0.380 ) |

**Supplementary Table 6:**
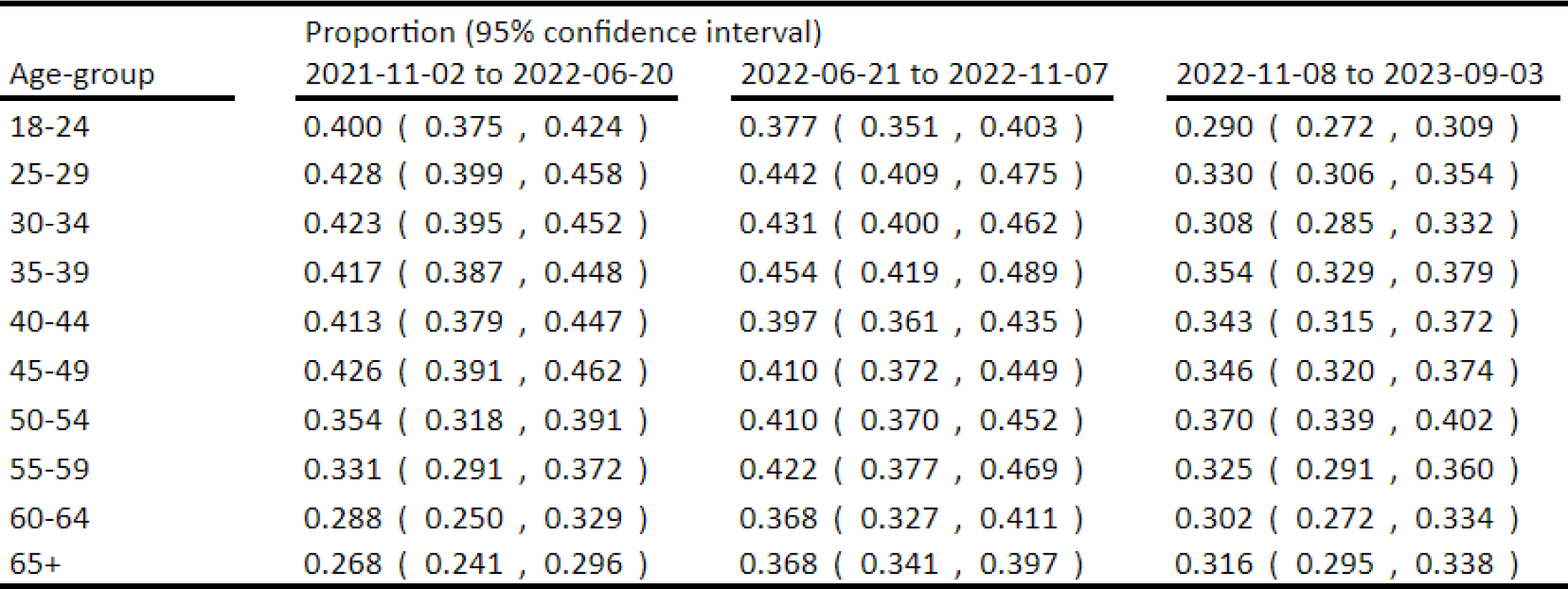
The probability of testing due to symptoms by age-group. The average (over three separate periods of the study) probability of an individual (by age-group) reporting testing for COVID-19 (with the presence of symptoms given as a reason for testing) given they were exhibiting at least one core symptom.

**Supplementary Table 7:** The probability of a test being positive. The average (over the entire study duration for which data was available: 8 February 2022 onwards) probability of a PCR test and **RAT** being positive by the symptom status of the individuals testing.

| Symptoms | Proportion of tests positive (95% confidence interval) |  |
| --- | --- | --- |
|  | PCR tests | RAT |
| at least one symptom | 0.364 ( 0.352 , 0.376 ) | 0.155 ( 0.150 , 0.161 ) |
| at least one core symptom | 0.403 ( 0.389 , 0.418 ) | 0.212 ( 0.205 , 0.220 ) |
| at least two core symptoms | 0.465 ( 0.445 , 0.486 ) | 0.312 ( 0.301 , 0.324 ) |
| at least fever and cough | 0.546 ( 0.513 , 0.578 ) | 0.458 ( 0.437 , 0.479 ) |

## Notes

### Competing Interest Statement

The authors have declared no competing interest.

### Author Declarations

The Human Research Ethics Committee of the University of Melbourne gave ethical approval for this work (reference number 2023-26949-40340-2)

